# Evaluation of a Multiplexed Immunoassay for Assessing Long-Term Humoral Immunity to Monkeypox virus infection and Orthopoxvirus Vaccination

**DOI:** 10.1101/2024.05.30.24308119

**Authors:** Bethany Hicks, Scott Jones, Helen Callaby, Daniel Bailey, Claire Gordon, Tommy Rampling, Catherine Houlihan, Ezra Linley, Simon Tonge, Clarissa Oeser, Rachael Jones, Marcus Pond, Ravi Mehta, Deborah Wright, Bassam Hallis, Cathy Rowe, Ashley Otter

## Abstract

In the summer of 2022, a large outbreak of Monkeypox virus (MPXV) cases occurred globally. By December 2022, a total of 3,582 Mpox cases had been confirmed within the UK. As a result, the Modified Vaccinia Ankara-Bavarian Nordic (“IMVANEX”) vaccine was offered to high-risk groups to protect against the spread of the virus. This outbreak led to the development of multiple serological assays to aid the current understanding of Mpox immunology. This study assessed the performance of a multiplexed solid-phase electrochemiluminescence (Meso Scale Discovery (MSD)) immunoassay for simultaneous detection of antibodies against MPXV A29, A35, B6, E8, and M1 antigens, along with the corresponding Vaccina Virus (VACV) homologues A27, A33, B5, D8, and L1. Sensitivity and specificity were evaluated with paediatric negatives (n=215), pre- and post-IMVANEX vaccinated (n=80) and MPXV (2022 Clade IIb outbreak, n=39) infected serum samples. The overall Orthopoxvirus multiplex assay demonstrated high specificity ranging from 75.68% (CI: 69.01-81.29) - 95.98% (CI:92.54-97.87) and sensitivity from 62.11% (CI:52.06-71.21) - 98.59% (CI:92.44% - 99.93%) depending on the Orthopoxvirus antigen, either used singularly or combined. Additionally, preferential binding was observed between Mpox-infected individuals and MPXV antigens, whilst vaccinated individuals exhibited increased binding to VACV antigens. These results highlight the differential binding patterns between antigen homologues in closely related viruses. Using this assay, we show that the Orthopoxvirus MSD assay is highly sensitive in detecting IgG titres for vaccinated sera ≥24-days post dose one and ≥14-days post dose two for all antigens within the assay except for MPXV A29 and VACV A27. A similar trend was observed with convalescent sera, although differing antigens demonstrate stronger sensitivities. Overall, this assay has the capability to accurately assess antibody titres for multiple relevant MPXV and VACV antigens post infection and post vaccination, demonstrating its utility in understanding immune responses to Orthopox viruses in current and future outbreaks, and assessing the immunogenicity of new generation Orthopox and Mpox-specific vaccinations.

## Introduction

MPXV is a zoonotic Orthopoxvirus (OPXV) characterised as a large dsDNA virus with a genome encoding over 200 different proteins^1^. This complex antigenic profile poses challenges with serological discrimination due to increased cross-reactivity of surface proteins. Furthermore, MPXV has two closely related clades; Clade I (formerly Congo Basin), and Clade II (formerly West African)^2^, whereby Clade IIb was the primary clade in the recent 2022 Mpox outbreak in various non-endemic countries^3–5^.

Vaccinia virus (VACV)^6,7^ has been essential in the development of various licensed smallpox vaccines^8^, which was successfully implemented to eradicate smallpox in the 1980s^9^. This eradication was possible due to the immunogenicity of VACV and its ability to provide cross-protection against additional Orthopoxviruses including Monkeypox Virus (MPXV), Cowpox virus (CPXV) and Variola virus (VARV)^10^. During the 2022 Mpox outbreak it was estimated that the efficacy of one IMVANEX vaccination dose against MPXV is ≥78%^11–14^.

It has been demonstrated that the recent second and third-generation Smallpox vaccines (ACAM2000 and IMVANEX, respectively) produce a similar antigenic response to MPXV infection^15^. This parallel immune response allows antibodies to bind to numerous MPXV and VACV antigens due to the high sequence homology in these proteins. Likewise, OPXV-elicited antibodies have increased cross-neutralising capability than other virus families^16^, although the research on discriminatory antigens and post-infection or vaccination immunity specific to MPXV antigens remain limited ^15,17,18^.

Surface proteins between VACV and MPXV-2022 exhibit approximately 94-98% sequence similarity^17^. During inter-host transmission, the mature virion is dominant where ∼25 membrane proteins are expressed, including MPXV A29 and E8 with VACV homologues A27 and D8 with an additional six proteins on the envelope (including MPXV A35 and M1)^18^ that play a dominant role in the intra-host transmission^19^. Due to the diverse number of immunodominant antigens, this initiated an increase in assay development by diverse groups and manufacturers to enable investigation of trends in serosurveillance, identification of MPXV cases, and evaluation of vaccine antibody waning necessitating booster doses^15,20,21^. However, many assays remain limited to single-plex, restricting the simultaneous evaluation of multiple antigens in a single well. Recognised for its high sensitivity multiplex capabilities, Meso Scale Discovery (MSD) developed an Orthopoxvirus assay that measures IgG antibody responses to five MPXV and five VACV immunodominant antigens. These range from cell adhesion through either binding chondroitin sulphate (MPXV E8 and VACV D8)^22^ or heparan sulphate (MPXV A29 and VACV A27)^23,24^, whereas MPXV B5 and VACV B6 are Extracellular Envelope virus (EEV)^25^ which have been identified to be a target for EEV-neutralising antibodies^26^.

A multiplex assay such as the Orthopoxvirus MSD panel described here enables higher-throughput screening of samples for quantifying antigen-specific antibodies. However, with next-generation Smallpox or Mpox-specific vaccines such as mRNA^27^, there is a need for more discrete immunology assays than using MVA or whole-virus serology assays. Here, we demonstrate the sensitivity and specificity of the MSD Orthopoxvirus assay that simultaneously measures IgG to five MPXV (A29, A35, B6, E8 and M1) and five VACV antigens (A27, A33, B5, D8 and L1).

## Materials and Methods

### Sample collection

#### Serum samples

Sera were obtained from individuals with an IMVANEX vaccination (n= 9 post-dose 1 (PD1), n*=*65 post dose 2 (PD2)), recruited from UKHSA Porton Down. Individuals were bled 24 days after dose one, then 14 days after dose two followed by approximate monthly bleeds up to 220 days after dose two. Anonymised Mpox convalescent serum (n*=*39) was obtained from PCR-confirmed infection from the 2022 outbreak (Clade IIb) from the Rare and Imported Pathogens Laboratory, Imperial College Healthcare NHS Trust and Chelsea and Westminster Hospital NHS Foundation Trust. To determine the assays specificity paediatric samples (n=215) from the UK Health Security Agency (UKHSA) Seroepidemiology Unit (SEU) were sourced as negative samples.

### Meso Scale Discovery (MSD) IgG Electrochemiluminescence immunoassay (ECLIA)

The MesoScale Discovery V-PLEX Orthopoxvirus Panel 1 (K15688U-2) electrochemiluminescence immunoassay (ECLIA) (MSD, Rockville, MD) was used to specifically measure IgG antibody binding to five MPXV (A29L, A35R, B6R, E8L and M1R) and five VACV (A27L, A33R, B5R, D8L and L1R) antigens in parallel in a 96-well 10 spot plate format. MSD assays were performed as per the manufacturer’s instructions. Briefly, 96-well plates were blocked with MSD Blocker A, following plate washing three times with 300 µl/well of PBS with Tween20 (0.01% final) using a Biotek 405 TS plate washer. Serum samples were diluted 1:1,000-10,000 in the manufacturers diluent buffer before being added to the MSD plate, along with the assay reference standard and controls. After incubation and a washing step, the detection antibody (MSD SULFO-TAG™ Anti-Human IgG Antibody) was added to each well. After further incubation and washing, MSD GOLD Read Buffer B was added, and plates read using the MESO QuickPlex™ SQ 120. A standard curve was established by fitting the signals from the standard using a 4-parameter logistic model. Concentrations of samples were determined from the electrochemiluminescence signals by back-fitting to the standard curve and multiplying by the dilution factor. Antibody concentrations are expressed in Arbitrary Units/ml (AU/ml). Kits were provided by MSD for evaluation purposes, with no involvement in the study design, result interpretation, or manuscript writing.

### Statistical analysis

Data analysis was performed using MSD Discovery Workbench 4.0 and GraphPad Prism version 9.2.0 (GraphPad, San Diego, CA).

### Ethics

IMVANEX Smallpox-vaccinated samples were obtained from individuals with written consent through UKHSA Research and Ethics Committee for assay validation. Mpox PCR-confirmed cases were granted approval for assay performance validation by the NHS Research Ethics Committee (reference 22/HRA/3321) as described by Otter *et al.*, 2023^15^.

## Results

### Evaluation of assay sensitivity and specificity for Orthopox vaccine and MPXV serology

Receiver operator curves (ROC) were plotted to determine the sensitivity and specificity using negatives against Mpox-infected and IMVANEX vaccinated samples for each antigen (**Table 1** and **Figure 1**). In addition to determine the area under the curve (AUC) to measure the accuracy of individual antigens. The AUC values were calculated based on the performance of the antigens in discriminating between negative and infected or vaccinated. Highest AUC values were observed for MPXV B6, MPXV E8, VACV A33, VACV B5 and VACV D8, with an AUC comparable for all antigens with a range of 0.76-0.99. **Table 1** indicates the positive/negative cut-offs for each antigen determined from the ROC analysis using a likelihood ratio (LR) calculation. MPXV A29 (62.11% (CI:52.06-71.21)) and its homologue VACV A27 (65.52% (CI:56.50-73.54)) were found to have much lower specificity and sensitivity compared to the other antigens when comparing negative samples with vaccinated and/or infected.

**Figure 1:**
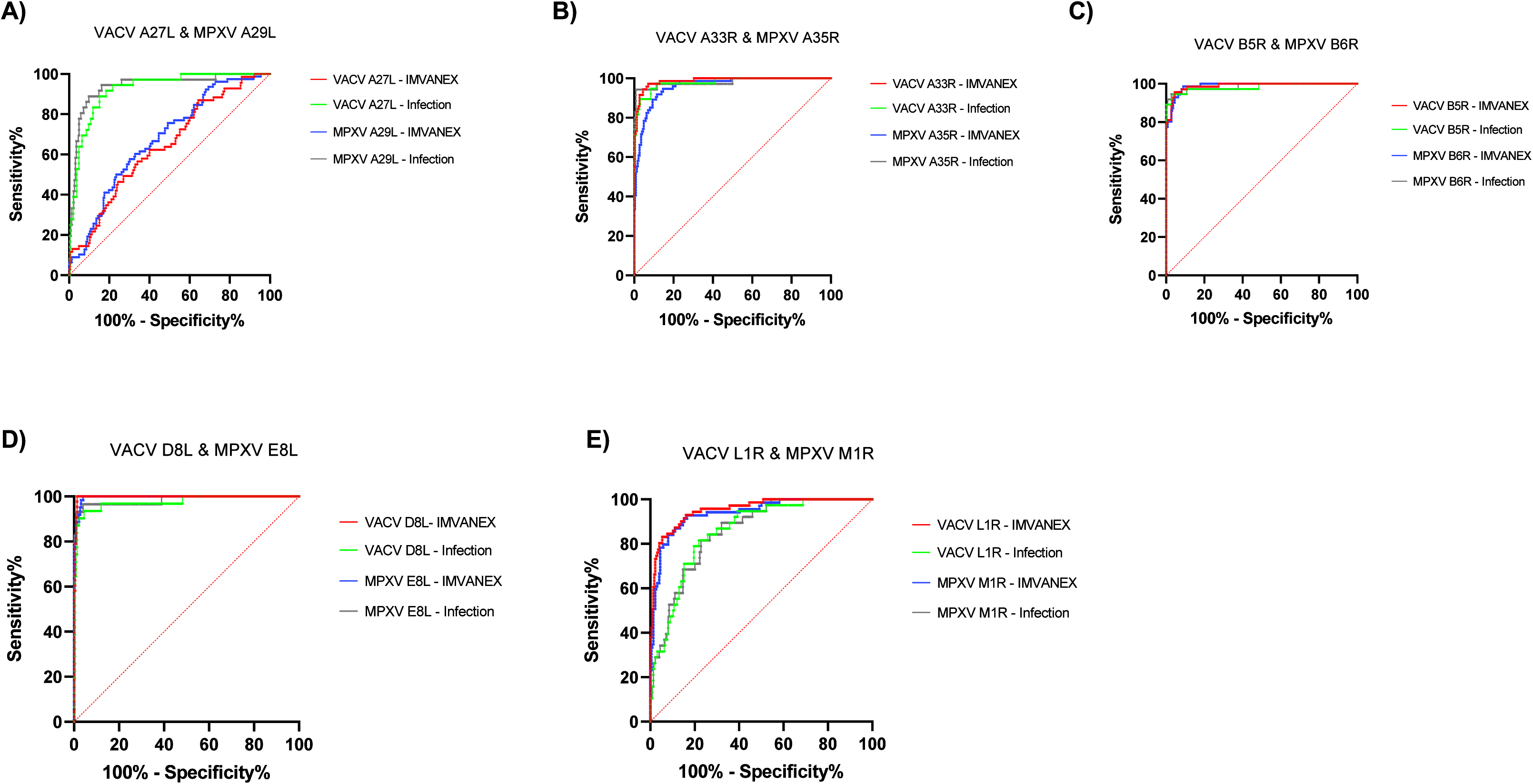
Receiver Operating Curves (ROC) for MPXV and VACV antigens, using negative samples as controls against positive samples as either post-Mpox infected (“Infection”) and/or post-IMVANEX vaccination (“IMVANEX”). ROC curves demonstrating the sensitivity versus 100%-specificity of MPXV antigens with post-vaccination (Blue) or post-infection (Black) and corresponding homologous VACV antigen with post-vaccination (Red) or post-infection (Green). Each homologous antigen pair from the MSD assay are included: (**A**) MPXV A29 and VACV A27, (**B**) MPXV A35 and VACV A33, (**C**) MPXV B5 and VACV B6, (**D**) MPXV E8 and VACV D8, and (**E**) MPXV M1 and VACV L1. Each solid line represents the ROC curve for the individual antigens and the dotted red line represents the random classifier. Negative samples (n=215) were compared against post Mpox-infection (n=39) and post IMVANEX vaccination samples (n=80).

**Table 1:** Comparative analysis of sensitivity and specificity for multiple MPXV and VACV antigens using ROC analysis. Sensitivity, specificity, and area under the curve (AUC) are stated for each individual antigen, as well as all 10 antigens combined based on the positivity score. Using paediatric negative (n=215) as controls, post Mpox-infection (n=39) and post IMVANEX vaccination (n=80) were compared.

| Antigen | Cut-off | Sensitivity % | 95% CI | Specificity % | 95% CI | Likelihood Ratio | AUC |
| --- | --- | --- | --- | --- | --- | --- | --- |
| MPXV A29L | > 2.358 | 62.11 | 52.06% to 71.21% | 82.43 | 76.89% to 86.87% | 3.54 | 0.784 |
| VACV A27L | > 2.112 | 65.52 | 56.50% to 73.54% | 75.68 | 69.01% to 81.29% | 2.69 | 0.757 |
| MPXV A35R | > 2.752 | 91.60 | 85.22% to 95.37% | 90.63 | 86.09% to 93.79% | 9.77 | 0.969 |
| VACV A33R | > 6.349 | 93.28 | 87.29% to 96.55% | 95.54 | 91.98% to 97.56% | 20.89 | 0.985 |
| MPXV B6R | > 2.666 | 94.12 | 88.35% to 97.12% | 95.52 | 91.94% to 97.55% | 20.99 | 0.989 |
| VACV B5R | > 6.151 | 95.61 | 90.14% to 98.11% | 95.52 | 91.94% to 97.55% | 21.32 | 0.987 |
| MPXV E8L | > 22.20 | 98.59 | 92.44% to 99.93% | 95.98 | 92.54% to 97.87% | 24.54 | 0.989 |
| VACV D8L | > 25.99 | 96.25 | 89.55% to 98.98% | 97.77 | 94.88% to 99.04% | 43.12 | 0.988 |
| MPXV M1R | > 11.19 | 83.90 | 76.22% to 89.44% | 84.82 | 79.54% to 88.93% | 5.53 | 0.913 |
| VACV L1R | > 10.37 | 85.83 | 78.48% to 90.96% | 83.93 | 78.55% to 88.16% | 5.34 | 0.924 |
| All Ten Antigens | > 4.50 | 92.92 | 86.65 % to 96.37% | 93.98 | 89.98% to 96.45% | 15.44 | 0.970 |

We next sought to assess ROC curves and therefore sensitivity and specificity for individual cohorts, separating vaccinated (IMVANEX dose 1 and 2 vaccinated) and MPXV convalescent sera (**Supplementary Table 1A & B**). Whilst specificity and sensitivity remained consistent across most antigens, MPXV A29 and VACV A27, showed higher sensitivity for MPXV convalescent sera in contrast to IMVANEX vaccinated sera sensitivity: 88.89% (CI:74.69-95.59) and 57.69% (CI:46.62-68.04), specificity: 90.09% (CI:85.45-93.36) and 69.82% (CI:63.49-75.48), respectively (**Table 1**, **Figure 1A**).

Conversely, MPXV M1 and VACV L1 demonstrated stronger sensitivity in detecting antibody responses in IMVANEX vaccinated sera compared to Mpox-infected sera, with a sensitivity of 91.30% (CI:82.30-95.95) and 81.58% (CI:66.58-90.78), and specificity: 84.82% (CI:79.54-88.93) and 77.23% (CI:71.31-82.24), respectfully (**Figure 1E**, **Table 1**).

### Utilising multiple antigens to determine Orthopox-positivity

As a multi-antigen assay, we sought to determine the ability to classify samples as positive based on the results to multiple antigens. Using the previously calculated cut-off values for each antigen (**Table 1**), we applied these to a panel of positive (vaccinated or infected) and negative samples, categorising samples as positive if they were greater than the respective cut-off values for each antigen. If a sample was above the cut off for each antigen, a value of one was assigned to it. The positivity count for each sample was assessed on a scale out of ten due to the 10 antigens. Using ROC analysis on negative versus positive samples, samples positive on 5 antigens resulted in a sensitivity of 92.92% (CI:86.65-96.37) and specificity of 93.98% (CI:89.98-96.45) (**Table 1**). Samples positive on greater than or equal to five antigens did not show an improvement in sensitivity or specificity, whilst less than five resulted in both decreases in sensitivity and specificity (**Supplementary Figure 2A**). This aligns with the observed trends in AUC values among IMVANEX-vaccinated and/or Mpox-infected individuals. For instance, the AUC ranges vary slightly across different cohorts, with values ranging from 0.734-0.944 for IMVANEX-vaccinated, 0.946-0.963 for Mpox-infected, and 0.807-0.950 for both cohorts combined. Notably, samples positive to five or more antigens consistently exhibit increased AUC values, reaching an equilibrium at approximately 0.97 (**Supplementary Figure 2A**).

### Comparative analysis of antibody titres to MPXV and VAVC antigens across cohorts

The evaluation of antibody titres across various cohorts revealed evident patterns for different antigens. Negative samples from paediatric samples predominantly demonstrated median antibody titres below the corresponding cut-off values (**Supplementary Table 2**). Most notably, the paediatric negative samples exhibited significantly (p<0.00001) lower antibody titres for all antigens except MPXV A29, A35, B6 and VACV B5 (**Figure 2**). Post-dose 1 (PD1) median antibody titres for all antigens were generally all positive; however, for MPXV A29, VACV A27, and MPXV A35, a number of results were borderline with the identified cut-off value for individual antigens (negative vs vaccination and Mpox-infected sera). Additionally, 14 days post-dose two (PD2) and further subsequent time points up to 84 days post dose two, significant increase in antibody levels was evident, displaying statistically significant (p<0.00001) differences from negatives for all antigens within the assay; MPXV A35, B6, E8, M1 and VACV A27 (p<0.0001), A33, B5, D8 and L1 (**Figure 2**).

**Figure 2:**
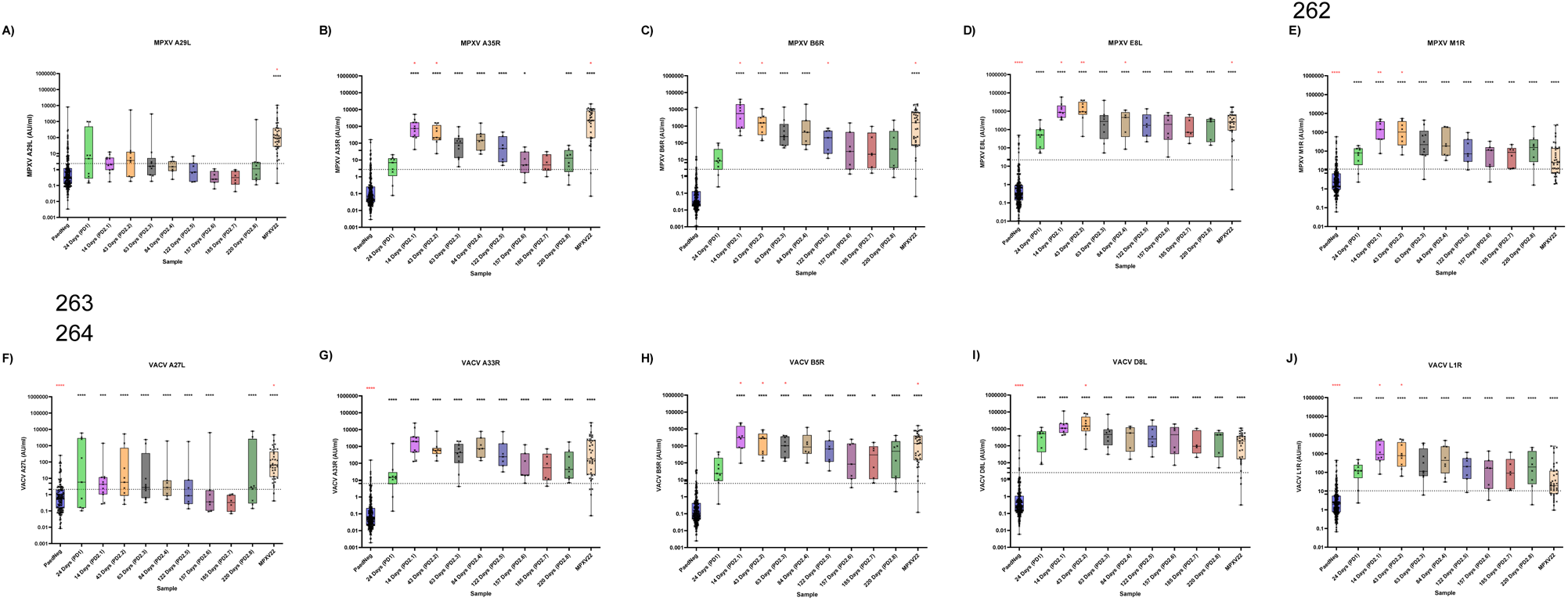
Antigen specific analysis of antibody titres across various cohorts including negative, IMVANEX vaccinated and Mpox-22 infected individuals. Statistical significance was determined between positive cohorts against paediatric negatives (black asterisk) or against post dose one (PD1) (red asterisk) using an unpaired T test where \**p* < 0.05, \*\**p* < 0.001, \*\*\**p* < 0.0001, and \*\*\*\**p* < 0.00001. The cut-off value for each antigen is represented by the dashed line. (**A**) MPXV A29, (**B**) MPXV A35, (**C**) MPXV B6, (**D**) MPXV E8, (**E**) MPXV M1, (**F**) VACV A27, (**G**) VACV A33, (**H**) VACV B5, (**I**) VACV D8 and (**J**) VACV L1.

Sera from vaccinated individuals were found to contain minimal antibodies directed towards MPXV A29 and VACV A27 antigens, with low levels of positivity and only 56% of positive samples exhibiting titres above the assigned cut-off value compared to other antigens, such as MPXV E8 and VACV D8 in which 100% of positive samples (vaccinated or infected) were found to be above the cut-off. Notably, positivity above the cut-off decreased from 63 days (PD2.3) and 122 days (PD2.5) post-second dose for MPXV A29 and VACV A27, with 0% positivity 157 days (PD2.6) and 185 days (PD2.7) post-second dose for MPXV A29 and for VACV A27, respectfully.

Conversely, 95% of Mpox-infected individuals are above the cut-off for MPXV A29 and VACV A27, with median antibody titres for these antigens at 96.07 AU/ml (CI:51.27-234.70) for MPXV A29 and 64.52 AU/ml (CI:32.15-145.50) for VACV A27. Compared to the average of all post-vaccination median values targeting these antigens, were markedly lower at 1.05 AU/ml (CI:0.60-2.04) and 2.02 AU/ml (CI:0.91-3.38), respectively (**Supplementary Table 2**).

### Analysis of serological reactivity to homologous MPXV/VACV proteins

In a comparative analysis of protein homologues, we demonstrate a notable trend in preferential binding between MPXV/VACV homologues (**Figure 3)**. We observed that sera from IMVANEX vaccinated individuals demonstrated preferential binding to VACV A33, B5 and D8, compared to the corresponding MPXV homologues A35, B6 and D8. Furthermore, we reveal contrasting antibody responses among MPXV convalescent sera compared to vaccine-induced sera. Mpox-infected individuals exhibit a stronger antibody response to MPXV A29 compared to its antigen homologue VACV A27 (**Figure 3A**). Mpox convalescent sera showed higher antibody levels (AU/ml) against MPXV A29 and VACV A27, with a majority (94%) of samples positive above the cut-off. Conversely, sera from vaccinated individuals demonstrated lower antibody responses, with over half (56%) of results falling below the corresponding cut-offs for these antigens (**Table 1**), parallel with the observations in **Figure 3A**.

**Figure 3:**
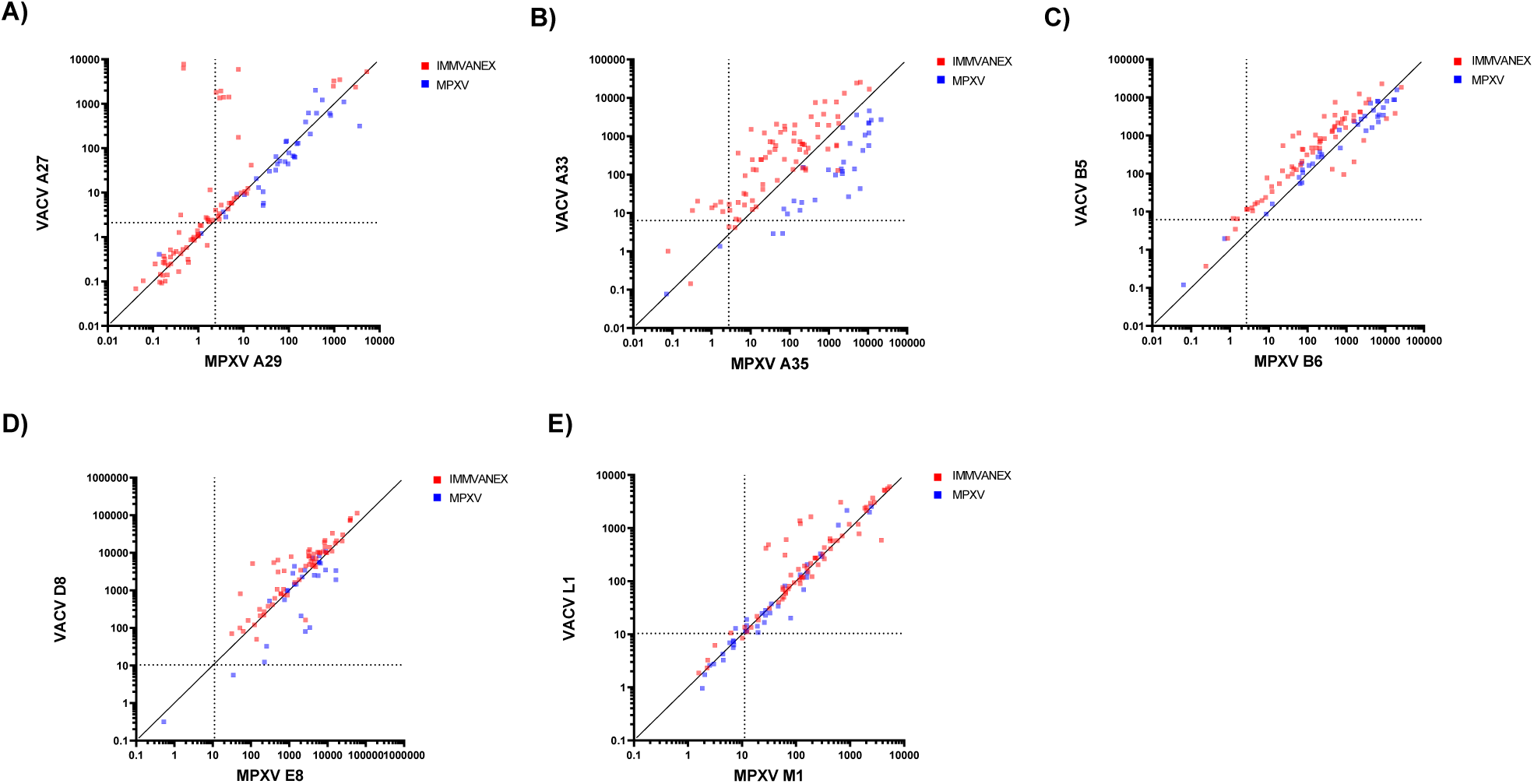
Correlation of post IMVANEX and MPXV convalescent sera to MPXV and VACV protein homologues: **A)** VACV A27 and MPXV A29 **B)** VACV A33 and MPXV A35 **C)** VACV B5 and MPXV B6 **D)** VACV D8 and MPXV E8 and **E)** VACV L1 and MPXV M1. The black dashed line response the corresponding cut-off for each antigen.

Conversely, MPXV M1 and VACV L1 antigens showed a reverse trend whereby IMVANEX-vaccinated individuals demonstrated preferential binding to VACV L1 rather than MPXV M1. Specifically, antibody positivity levels among Mpox-infected individuals were lower and border the cut-off value (24.64 AU/ml for MPXV M1 and 19.57 AU/ml for VACV L1, **Supplementary Table 2**). In contrast, vaccination-induced antibody responses post dose two displayed >90% positivity above the cut-off, with slightly lower positivity post dose one (78% and 89% against MPXV M1 and VACV L1, respectfully). Whereas Mpox-infected sera which exhibited antibody levels clustering around the cut-off, with 30% of samples falling below the threshold.

### Correlation between MXPV and VACV antigens

Using Pearson correlation, we wanted to determine how antibody binding results for each sample compare to one another for each VACV and MPXV antigen across Mpox-infected and IMVANEX vaccinated individuals. We observe that homologous proteins MPXV E8/VACV D8 and MPXV M1/VACV L1 showed the strongest correlation (**Figure 4, Supplementary Table 3**). In addition to the correlation observed between the homologous proteins, we observed that some antigens were strongly correlated to one another. In particular, MPXV A35/VACV A33 strongly correlated with VACV B5/MPXV B6, whilst VACV L1/MPXV M1 correlated to VACV A33, VACV D8/MPXV E8, and VACV B5/MPXV B6.

**Figure 4:**
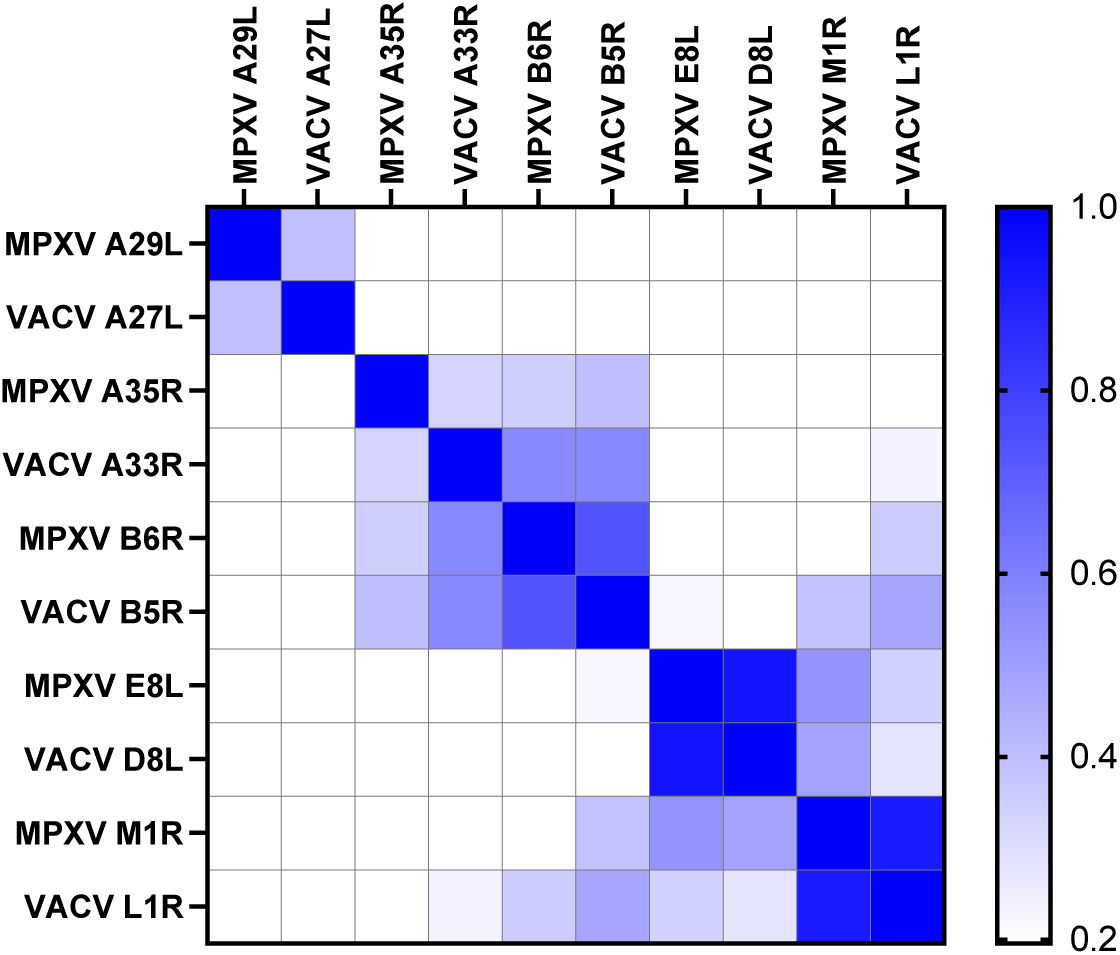
Two-tailed Pearson correlation analysis and heat map visualisation of significant correlations for antibody binding to each MPXV or VACV antigens for vaccinated and/or infected individuals. Blank cells represent non-significant (p ≥ 0.05) correlations.

## Discussion

In response to the global Mpox outbreak in 2022, a number of assays have been developed to assess both immune responses to MPXV infection and Smallpox vaccination^20,28^. Accurate assays for detecting MPXV and VAVC antibodies are critical for evaluating new vaccinations and for conducting serosurveillance studies to determine exposure to MPXV and those previously vaccinated, especially with ongoing Mpox infections in Europe, America and Asia, in addition to high-incidence countries such as DRC and Nigeria^29^.

This study aimed to evaluate the sensitivity and specificity of the MSD Orthopoxvirus 10-plex assay, coated with immunodominant antigens MPXV M1, A35, B6 and A29 which have previously been identified to be highly immunogenic^21,30–32^. We applied this assay for the subsequent analysis of antibody titres across both infected and IMVANEX (“JYNNEOS”) vaccination cohorts to provide an understanding of the humoral immune responses to both MPXV and VACV antigens. We have demonstrated this assay exhibits a high level of specificity and sensitivity in detecting antibodies against MPXV and VACV antigens, revealing antigen specific immune responses post infection and/or vaccination.

### Orthopoxvirus MSD panel demonstrates high sensitivity and specificity in detecting Orthopoxvirus antibodies

Individual median fluorescence intensity (MFI) cut-offs were determined for each antigen using ROC analysis by comparing negatives against IMVANEX vaccinated and Mpox-infected (**Supplementary Figure 1**). The sensitivity and specificity varied across the ten antigens, with corresponding area under the curves (AUCs) ranging from 0.757 (VACV A27) to 0.989 (MPXV E8). Certain antigens, such as MPXV A35, B6 and E8, as well as VACV A33, B5 and D8, demonstrated high accuracy levels (AUC values >0.96). This indicates the potential for these antigens in distinguishing between vaccinated and/or Mpox-infected individuals to negative individuals. However, when separating positive samples to either post Mpox-infected or post IMMVANEX vaccinated, it was evident specific antigens (MPXV A29, M1 and VACV A27 and L1) have differences in sensitivity and specificity values (**Figure 1**). While both MPXV M1 and VACV L1 exhibit AUC values >0.9, there is a small discrepancy between post IMVANEX vaccination and post Mpox-infection towards the sensitivity and specificity for these antigens. An ROC analysis evaluating paediatric negatives to vaccinated sera reveals a higher sensitivity (91.3 % CI:82.30-95.95) compared to those infected with Mpox (81% CI:66.58-90.78) and vaccinated and Mpox-infected individuals combined (83.9% CI:76.22-89.44) (**Table 1, Supplementary Table 1**). This demonstrates that individuals vaccinated with IMVANEX generate a higher M1 antibody response compared to those infected, consistent with findings from previous studies^28^. Additionally, it has been observed that mRNA vaccines (VGPox1 and VGPox2, both encoding a fusion protein containing A35R and M1R) induce a rapid and robust production of MPXV M1 and A35 specific antibodies seven days after vaccination, indicating a more effective immune response to MPXV M1 compared to natural infection which induced humoral immunity against other antigens with non-significant levels of M1-specific antibodies seven days post exposure^33^.

Similarly, MPXV A29 and VACV A27 demonstrated lower specificity and sensitivity, emphasising potential limitations in distinguishing between positive and negative samples for these antigens, although they had AUCs of 0.78 and 0.76, respectively, indicating adequate accuracy. Likewise, Yates *et al.*^20^ demonstrated that during the development of a serological assay MPXV antigen A29 and its homologue VACV A27 had a low sensitivity and specificity with the AUC <0.80, which showed similarities with the antigens in this multiplexed assay. Interestingly, with vaccinated samples removed from the ROC analysis, this increases the specificity (89% (CI:74.69-95.59)) and specificity (90% (CI:85.45-93.36)), and likewise for VACV A27 sensitivity (84% (CI:78.39-88.86)) and specificity (88.89% (CL:74.69%-95.59)) increases (**Figure 1A**). This suggests there could be variation in the immune response between convalescent individuals and those who received the IMVANEX vaccination for this antigen. It has been identified that MPXV A29 is the main antigen used in immunoassays for MPXV due to its location on the envelope^34^, this suggests that individuals infected with MPXV would likely generate a higher antibody response to MPXV A29 compared to those vaccinated, as seen in this assay **(Figure 2A)**. Likewise, MPXV A29 (along with additional MPXV proteins A35, M1 and B6) have been identified to generate a CD4+ T cell cellular response and are one of the primary targets of neutralising antibodies^21,35^.

### Determining optimal assay cut-offs for detecting broad and specific Orthopoxvirus antibodies

This assay shows that samples positive to 1 - 4 antigens have a lower specificity and sensitivity compared to ≥5 antigens (**Supplementary Figure 2A**). The cut-off of ≥5 antigens demonstrate a high sensitivity (93% CI:86.65-96.37) and specificity (94% CI:89.98-96.45), whilst evaluating different cohorts (convalescent versus vaccinated) to assess if an individual is positive to Orthopox antibodies. Whilst a limited number of commercial assays exist for Mpox serology, the MSD multiplex assays offers a unique opportunity to assess antibody titres to individual antigens but also through our classification method described here can be used for inferring the Orthopox-specific immune status. With the established assay cut-off, this can be used to classify individuals as positive or negative for Orthopoxvirus antibodies should they be above the identified cut off for ≥5 antigens. This established assay offers a promising tool for both diagnostic and serosurveillance applications, thereby enhancing further serological research around Mpox in an easy-to-use format. A number of additional antigens could be included in future assay developments, such as MPXV A27 which have been shown to be differential for vaccinated from infected^15,36^.

### Orthopoxvirus-specific IgG antibody dynamics vary by antigen

Across the MPXV/VACV antigens we observe that Smallpox-vaccination (IMVANEX) induces a similar antibody response to those infected with MPXV Clade IIb. Due to their cross protection (as well as high genetic and antigenic homology), Smallpox vaccines have been used to prevent and reduce the severity of Mpox^13^. Although currently the Orthopoxvirus MSD plates are limited to five antigen homologues, future generations of Orthopoxvirus vaccines are likely to be antigen specific as with the development of a number of multivalent mRNA vaccines^27,37^, highlighting the necessity for multiplex assays for Orthopoxvirus immunity testing.

Whilst the Mpox genome has >200 proteins, the MSD assay is limited to 10 antigens in total (five of each MPXV and VACV homologous proteins), therefore other antigens capable of generating an immune response could be undetected in this assay. For instance, we previously demonstrated MPXV A27 as a potential discriminatory antigen between infected and vaccinated individuals due the truncation of the A27L gene in MVA-BN (IMVANEX)^15^. In addition, we demonstrated MPXV B2 as a primary immunodominant antigen after one IMVANEX dose^15^. The inclusion of additional antigens such as these in newer generation serological assays are paramount to identifying antigens involved in protective immune responses against Orthopoxviruses, as well as discriminating between vaccine- and infection-derived immunity, both for vaccine and serosurveillance studies.

Similarly, although the MVA-BN vaccine primarily induced antibodies specific to MVA and VACV, there is a limited correlation between MVA and MPXV neutralising antibodies^38^. This suggests antigenic differences among Orthopoxviruses, an area that should be investigated further to evaluate vaccine immunogenicity and cross-protection. Notably, this study only focused on Mpox Clade IIb, although it has been observed there is no difference in antigen recognition between Clade IIa^15^, there are mutational differences observed between variants^39^ which could lead to divergent immune responses, an area to be explored further. Especially with the continued spread of Clade I Mpox evidenced by recent outbreaks in the Democratic Republic of Congo^29^.

Analysis of antibody titres revealed distinct patterns across cohorts, providing essential information on the dynamics of the humoral immune response. Negative samples predominantly exhibited antibody titres below the cut-off values for all antigens, indicating either undetectable immune responses, or in those with low level reactivity being presumed cross-reactive antibodies and background noise. IMVANEX-vaccinated individuals post-dose one displayed an initial low, yet detectable and significantly higher (p=<0.00001) immune response against MPXV E8, M1, and VACV A27, A33, D8 and L1 antigens compared to negatives (**Figure 2**). However, the lack of antibody responses indicated here does not suggest lack of protection, with IMVANEX vaccine efficacy after one dose assessed as ≥78%^11–14^.

A significant increase in antibody titres is observed for MPXV A35 (p=0.021), B6 (p=0.026), E8 (p=0.022) and VACV B5 (p=0.042), L1 (p=0.022) in relation to PD1, 14-days from administration of the second dose (**Figure 2**), with VACV D8 (p=0.038) demonstrating significant increases 43-days post second vaccination, suggesting a necessity for a second dose, similarly observed in other (singleplex) assays^15,16^. Although it is shown antibodies are present, whether these antibodies provide protection remains unanswered, emphasising the need for further investigation into correlates of protection and antigen-specific responses on overall immunity.

Nevertheless, a subsequent decrease in antibody titres was observed ∼84 days post-dose 2 (PD2.4, **Figure 2**), suggesting a waning of the humoral immune response to MPXV and VACV antigens. A similar trend that we previously demonstrated with individual MPXV and VACV antigen ELISAs^28^. The majority of samples remain above the designated cut-off defined in this study up to 220 days post-dose two. After this, median antibody titres decline for each antigen and are non-significantly different to post dose one responses. MPXV A35 and B6 demonstrate the most pronounced waning of immune responses post two doses of vaccination, specifically, antibody titres wane to levels comparable to negative samples >185 days for MPXV A35 (PD2.7, **Figure 2B**) and >122 days for MPXV B6 (PD2.5, **Figure 2C**). The continued waning of antigen-specific antibodies is being sought from ongoing longitudinal investigations, using a panel of 12 MPXV-specific antigens, some of which can be assessed through this MSD assay.

### Correlation between IMVANEX and Mpox-infected individuals against MPXV/VACV homologous proteins

As previously shown^15^ when comparing antigen homologues IMVANEX vaccinated sera showed preferential binding to VACV A33, B5 and D8 and MPXV convalescent sera to MPXV A35, B6 and E8, respectively. MPXV A35 and VACV A33 had the most pronounced differentiation (**Figure 3**) which corresponds to findings observed in other singleplex and multiplex ELISAs^15,36^, highlighting that the type of antigen exposure influences the subsequent antibody binding preference. All antigens within this assay are MPXV and VACV homologous proteins, which could provide additional mechanisms for differentiating between convalescent and vaccine-derived antibodies. We observed that antibodies against MPXV A29 and VACV A27 were generally higher in Mpox convalescent samples than those post-IMVANEX vaccination. MPXV A29 is highly conserved between MPXV and VACV^40^ with a 93.6% sequence identity with VACV A27 homologue,^41^ so the observation of a decreased humoral immune response to MPXV A29 is not known and warrants further investigation. Instances of truncated proteins (e.g., MPXV A27) have been observed in the MVA-BN strain, suggesting something similar may have occurred which could aid further aid in the differentiation between infected, historically vaccinated and recently vaccinated. However, whilst there are reduced antibody titres, these antigens activate cellular immunity and produce neutralising antibodies, providing a level of protection^21^. Further investigations into MPXV A29 and VACV A27 could provide a valuable insight into immunity dynamics.

Conversely, with MPXV M1 and VACV L1 we observe antibody responses to these antigens more evidently in those receiving the IMVANEX vaccine (**Figure 2**). MPXV M1 has been identified as an important immunogen^42^ and has 98.8% sequence homology with VACV L1^41^, suggesting that the robust antibody response of M1 should be mirrored in the homologous VACV protein. Due to the robust antibody and T-cell responses with MPXV M1^27^, this has been included in candidate multivalent mRNA Orthopoxvirus vaccine along with MPXV A35, B6 and H3. We have similarly confirmed the observation of M1 (and therefore L1) antibodies being vaccination specific^15^, warranting further investigation as to the mechanism as to why these are vaccination specific. Finally, using Pearson correlation we observed the relationship between MPXV/VACV antigens. All antigens are significantly correlated to their corresponding homologue, with MPXV E8/VACV D8 exhibiting the strongest correlation (**Figure 4, Supplementary Table 3**). Additionally, we observed correlations between certain antigens such as MPXV A35/VACV A33 correlated with MPXV B6/VACV B5, as well as MPXV M1/VACV L1 correlated to VACV A33, MPXV E8/VACV D8, and MPXV B6/VACV B5. These correlations primarily involve immunodominant antigens^15,21^, suggesting their importance and the relationship between different antigens in the overall immune response to infection and vaccination.

## Conclusion

Here we demonstrate the MSD Orthopoxvirus assay is highly sensitive, specific, and reproducible for the detection of Orthopox-specific antibodies. This assay provides a valuable tool for serosurveillance and in the assessment of new generation Orthopox vaccine candidates as well as understanding Orthopoxvirus immunity. This assay provides benefits over current singleplex assays in its ability to test multiple antigens simultaneously, with opportunities for comprehensive studies and deeper insights into the ongoing spread of Mpox worldwide.

## Data Availability

All data produced in the present study are available upon reasonable request to the authors

---

**Figure 1:**
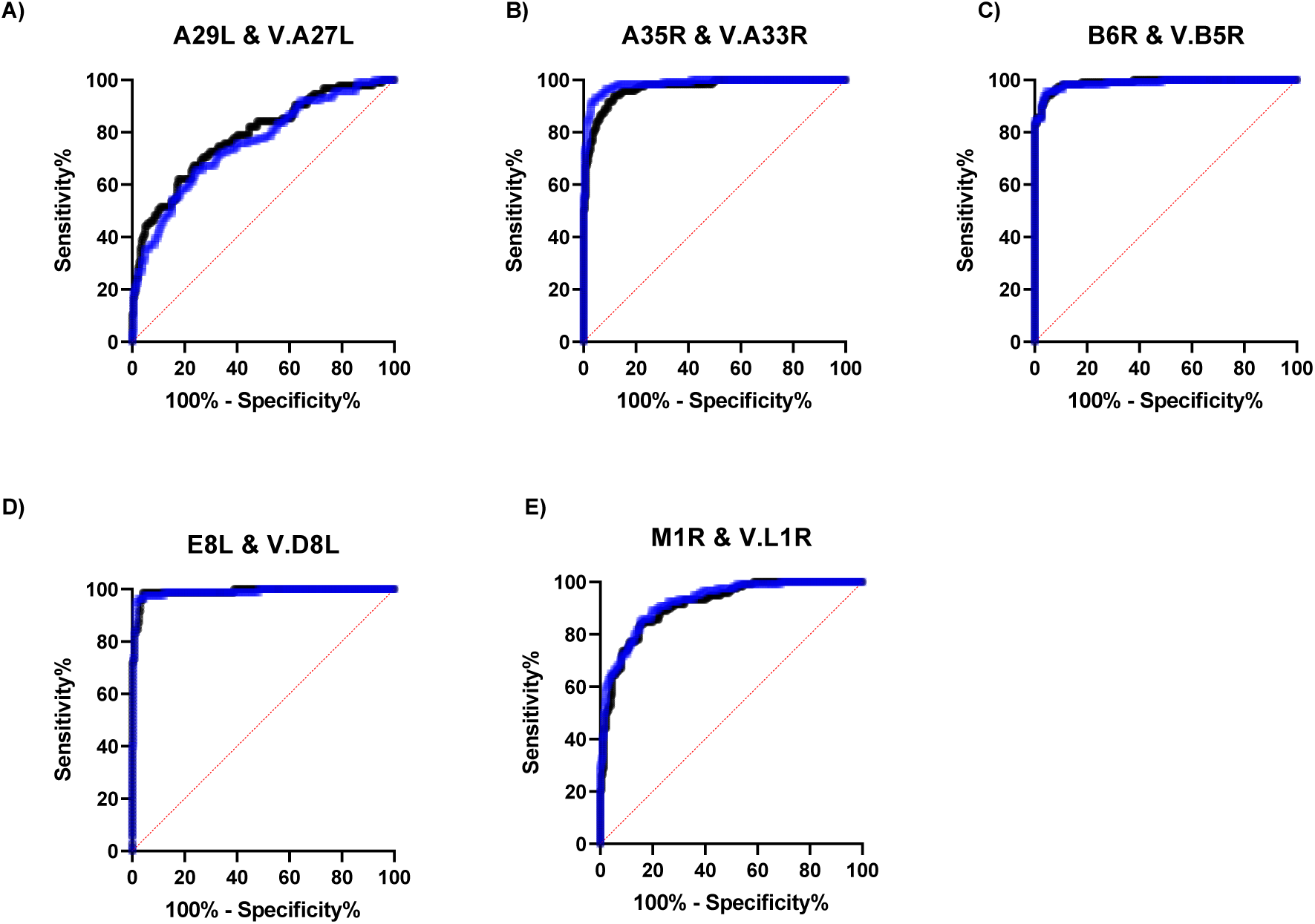
Receiver Operating Curves (ROC) for MPXV (black) and VACV (blue) homologous proteins. Evaluating the sensitivity and specificity of homologues with negative (n=215) versus Mpox-infected (n=39) and IMVANEX vaccinated (n=80) individuals. **A)** MPXV A29 & VAC A27, **B)** MPXV A35 & VACV A33, **C)** MPXV B6 & VACV B5, **D)** MPXV E8 & D8 and **E)** MPXV M1 & VACV L. Each solid line represents the ROC for individual antigens, while the dashed red line is a random classifier.

**Figure 2:**
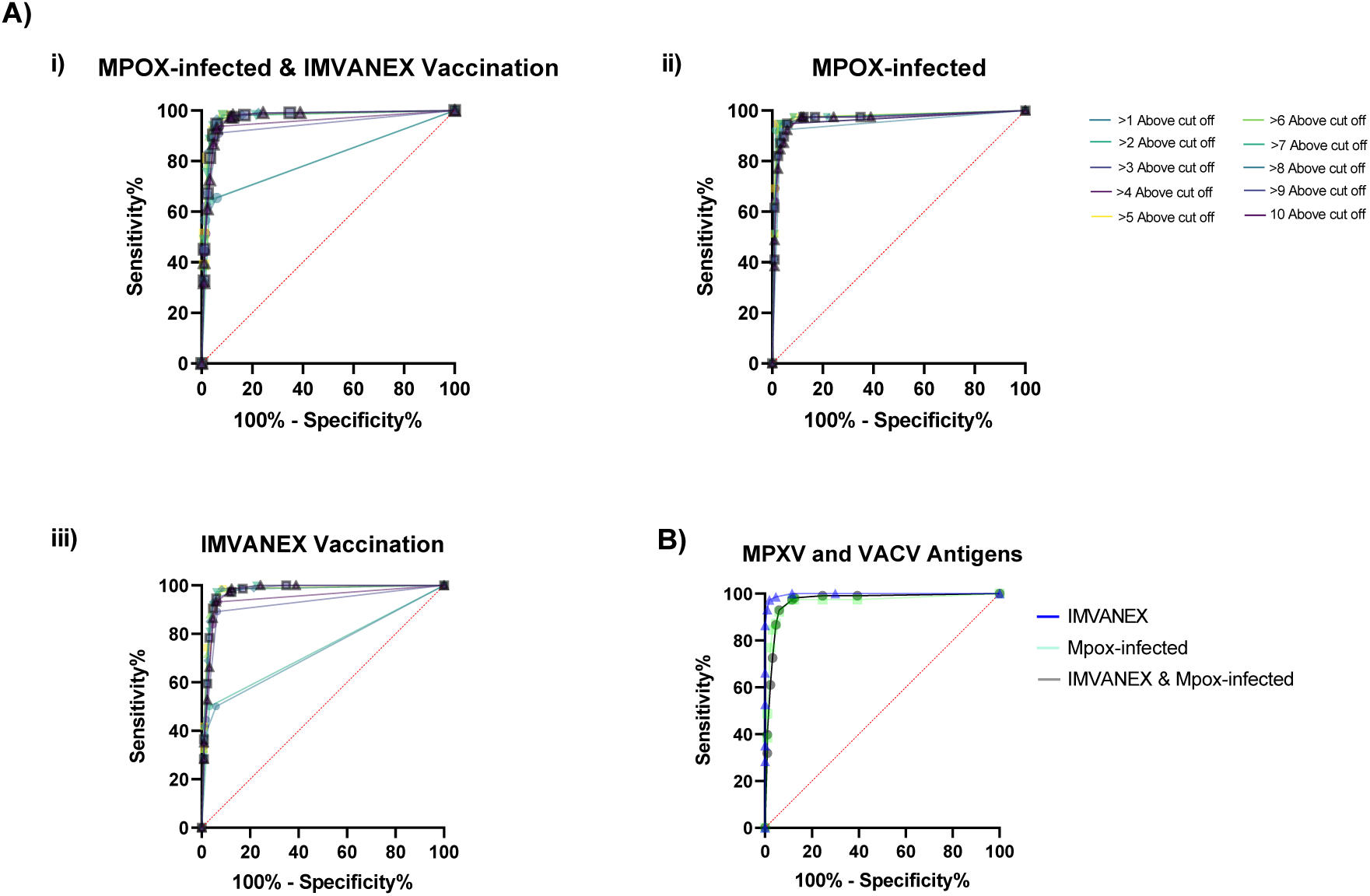
Combined antigen ROC curves. **A)** Comparing negative versus either **(i.)** Mpox-infected & IMVANEX infected, **(ii.)** Mpox-infected or **(iii.)** IMVANEX vaccinated. Evaluating antigen positivity across a spectrum of one to ten antigens relative to cut-off values detailed in Table 1. **B)** ROC curve for all ten antigens based on positivity scores, for IMVANEX vaccinated, Mpox-infected and both combined.

**Table 1:** Summary of ROC curves and individual antigen cut offs. **A)** Mpox-infected and **B)** IMVANEX vaccinated. Cut-offs were generated but comparing all positive samples (either Mpox-infected (n=39) or IMVANEX vaccinated (n=80) with all negative samples to each MPXV/VACV antigen by ROC analysis.

| Antigen | Cut off | Sensitivity % | 95% CI | Specificity % | 95% CI | Likelihood Ratio | AUC |
| --- | --- | --- | --- | --- | --- | --- | --- |
| **A) Mpox-Infected** |  |  |  |  |  |  |  |
| MPXV A29L (NEGS Vs MPXV) | > 6.976 | 88.89 | 74.69% to 95.59% | 90.09 | 85.45% to 93.36% | 8.97 | 0.938 |
| MPXV A35R (NEGS Vs MPXV) | > 37.09 | 94.29 | 81.39% to 98.98% | 99.11 | 96.80% to 99.84% | 105.61 | 0.981 |
| MPXV B6R (NEGS Vs MPXV) | > 6.706 | 94.59 | 82.30% to 99.04% | 97.31 | 94.26% to 98.76% | 35.16 | 0.986 |
| MPXV E8L (NEGS Vs MPXV) | > 23.83 | 94.44 | 74.24% to 99.72% | 95.98 | 92.54% to 97.87% | 23.51 | 0.975 |
| MPXV M1R (NEGS Vs MPXV) | > 6.852 | 81.58 | 66.58% to 90.78% | 77.23 | 71.31% to 82.24% | 3.58 | 0.856 |
| VACV A27L (NEGS Vs MPXV) | > 5.014 | 88.89 | 74.69% to 95.59% | 84.32 | 78.39% to 88.86% | 5.67 | 0.923 |
| VACV A33R (NEGS Vs MPXV) | > 9.251 | 89.47 | 75.87% to 95.83% | 96.88 | 93.69% to 98.48% | 28.63 | 0.989 |
| VACV B5R (NEGS Vs MPXV) | > 8.307 | 94.59 | 82.30% to 99.04% | 96.41 | 93.08% to 98.17% | 26.37 | 0.982 |
| VACV D8L (NEGS Vs MPXV) | > 11.84 | 93.55 | 79.28% to 98.85% | 95.54 | 91.98% to 97.56% | 20.95 | 0.974 |
| **B) IMVANEX Vaccinated** |  |  |  |  |  |  |  |
| MPXV A29L (NEGS Vs IMVANEX) | > 0.9492 | 57.69 | 46.62% to 68.04% | 69.82 | 63.49% to 75.48% | 1.91 | 0.6756 |
| MPXV A35R (NEGS Vs IMVANEX) | > 2.752 | 89.19 | 80.09% to 94.42% | 90.63 | 86.09% to 93.79% | 9.51 | 0.9601 |
| MPXV B6R (NEGS Vs IMVANEX) | > 1.112 | 97.18 | 90.30% to 99.50% | 91.93 | 87.60% to 94.83% | 12.04 | 0.9887 |
| MPXV E8L (NEGS Vs IMVANEX) | > 22.20 | 100 | 92.29% to 100.0% | 95.98 | 92.54% to 97.87% | 24.89 | 0.9943 |
| MPXV M1R (NEGS Vs IMVANEX) | > 11.19 | 91.3 | 82.30% to 95.95% | 84.82 | 79.54% to 88.93% | 6.02 | 0.9391 |
| VACV A27L (NEGS Vs IMVANEX) | > 2.112 | 46.38 | 35.11% to 58.02% | 75.68 | 69.01% to 81.29% | 1.91 | 0.6413 |
| VACV A33R (NEGS Vs IMVANEX) | > 6.349 | 94.37 | 86.39% to 97.79% | 95.54 | 91.98% to 97.56% | 21.14 | 0.9865 |
| VACV B5R (NEGS Vs IMVANEX) | > 6.151 | 95.65 | 87.98% to 98.81% | 95.52 | 91.94% to 97.55% | 21.33 | 0.9885 |
| VACV D8L (NEGS Vs IMVANEX) | > 48.38 | 100 | 90.11% to 100.0% | 98.66 | 96.14% to 99.63% | 74.67 | 0.9968 |
| VACV L1R (NEGS Vs IMVANEX) | > 10.37 | 92.96 | 84.55% to 96.95% | 83.93 | 78.55% to 88.16% | 5.78 | 0.9545 |

**Table 2:**
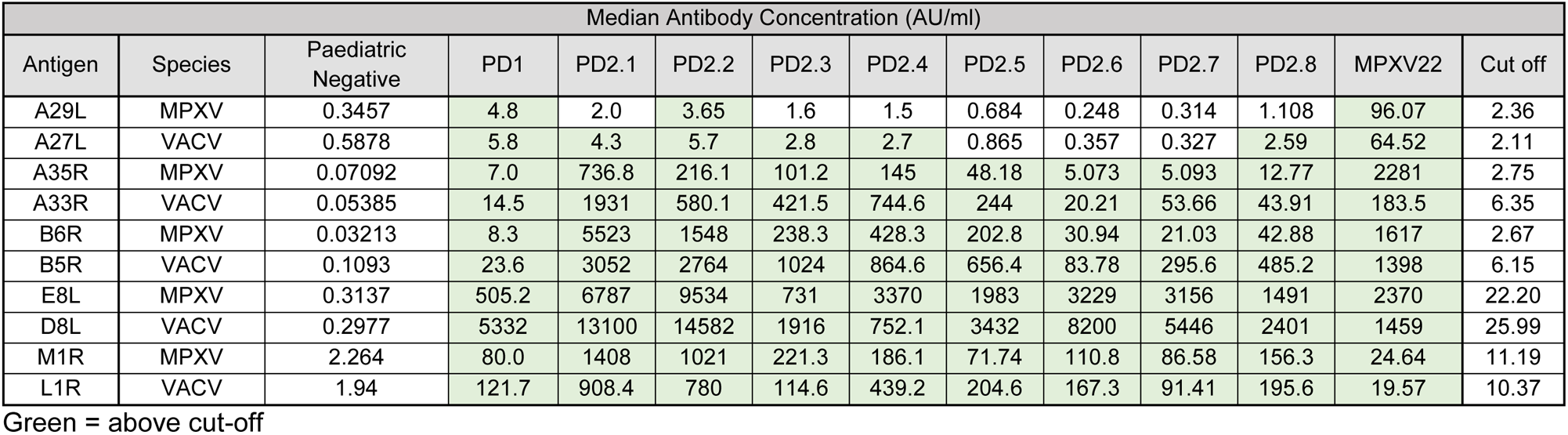
Median antibody concentrations (AU/ml) for all MPXV and VACV antigens. For paediatric negatives (n=215), 24 days post-dose one (PD1), 14 days post-dose two (PD2.1), with following longitudinal timepoints: 43 days (PD2.2), 43 days (PD2.2), 63 days (PD2.3), 84 days (PD2.4), 122 days (PD2.5), 157 days (PD2.6), 185 days (PD2.7), 220 days (PD2.8) and Mpox-infected (n=39). The corresponding cut-off for each antigen was assigned using ROC analysis. Cells highlighted in green show the median concentrations to be above the assigned cut-off.

| Median Antibody Concentration (AU/ml) |  |  |  |  |  |  |  |  |  |  |  |  |  |
| --- | --- | --- | --- | --- | --- | --- | --- | --- | --- | --- | --- | --- | --- |
| Antigen | Species | Paediatric Negative | PD1 | PD2.1 | PD2.2 | PD2.3 | PD2.4 | PD2.5 | PD2.6 | PD2.7 | PD2.8 | MPXV22 | Cut off |
| A29L | MPXV | 0.3457 | 4.8 | 2.0 | 3.65 | 1.6 | 1.5 | 0.684 | 0.248 | 0.314 | 1.108 | 96.07 | 2.36 |
| A27L | VACV | 0.5878 | 5.8 | 4.3 | 5.7 | 2.8 | 2.7 | 0.865 | 0.357 | 0.327 | 2.59 | 64.52 | 2.11 |
| A35R | MPXV | 0.07092 | 7.0 | 736.8 | 216.1 | 101.2 | 145 | 48.18 | 5.073 | 5.093 | 12.77 | 2281 | 2.75 |
| A33R | VACV | 0.05385 | 14.5 | 1931 | 580.1 | 421.5 | 744.6 | 244 | 20.21 | 53.66 | 43.91 | 183.5 | 6.35 |
| B6R | MPXV | 0.03213 | 8.3 | 5523 | 1548 | 238.3 | 428.3 | 202.8 | 30.94 | 21.03 | 42.88 | 1617 | 2.67 |
| B5R | VACV | 0.1093 | 23.6 | 3052 | 2764 | 1024 | 864.6 | 656.4 | 83.78 | 295.6 | 485.2 | 1398 | 6.15 |
| E8L | MPXV | 0.3137 | 505.2 | 6787 | 9534 | 731 | 3370 | 1983 | 3229 | 3156 | 1491 | 2370 | 22.20 |
| D8L | VACV | 0.2977 | 5332 | 13100 | 14582 | 1916 | 752.1 | 3432 | 8200 | 5446 | 2401 | 1459 | 25.99 |
| M1R | MPXV | 2.264 | 80.0 | 1408 | 1021 | 221.3 | 186.1 | 71.74 | 110.8 | 86.58 | 156.3 | 24.64 | 11.19 |
| L1R | VACV | 1.94 | 121.7 | 908.4 | 780 | 114.6 | 439.2 | 204.6 | 167.3 | 91.41 | 195.6 | 19.57 | 10.37 |

Green = above cut-off

**Supplementary Table 3:** Correlation between MPXV/VACV antigens using Pearson r. Significant values (p≤0.05) are indicated with corresponding p values.

|  | MPXV A29L | VACV A27L | MPXV A35R | VACV A33R | MPXV B6R | VACV B5R | MPXV E8L | VACV D8L | MPXV M1R | VACV L1R |
| --- | --- | --- | --- | --- | --- | --- | --- | --- | --- | --- |
| MPXV A29L | 1.000 | 0.393<br>($p=2.13E-05$) | 0.026 | -0.047 | 0.036 | 0.161 | 0.019 | -0.041 | 0.145 | 0.103 |
| VACV A27L | 0.393<br>($p=2.13E-05$) | 1.000 | -0.079 | -0.096 | -0.094 | -0.044 | -0.145 | -0.115 | -0.038 | -0.046 |
| MPXV A35R | 0.026 | -0.079 | 1.000 | 0.329<br>($p=0.0005$) | 0.346<br>($p=0.0003$) | 0.397<br>($p=3.25E-05$) | 0.044 | -0.110 | -0.130 | -0.094 |
| VACV A33R | -0.047 | -0.096 | 0.329<br>($p=0.0005$) | 1.000 | 0.568<br>($p=1.00E-10$) | 0.569<br>($p=1.93E-10$) | 0.148 | 0.086 | 0.033 | 0.232<br>($p=0.0157$) |
| MPXV B6R | 0.036 | -0.094 | 0.346<br>($p=0.0003$) | 0.568<br>($p=1.00E-10$) | 1.000 | 0.734<br>($p=3.76E-19$) | 0.191 | 0.018 | 0.195<br>($p=0.0457$) | 0.353<br>($p=0.0002$) |
| VACV B5R | 0.161 | -0.044 | 0.397<br>($p=3.25E-05$) | 0.569<br>($p=1.93E-10$) | 0.734<br>($p=3.76E-19$) | 1.000 | 0.223<br>($p=0.0370$) | 0.103 | 0.387<br>($p=5.48E-05$) | 0.474<br>($p=3.37E-07$) |
| MPXV E8L | 0.019 | -0.145 | 0.044 | 0.148 | 0.191 | 0.223<br>($p=0.0370$) | 1.000 | 0.94<br>($p=7.37E-43$) | 0.531<br>($p=1.22E-07$) | 0.339<br>($p=0.0012$) |
| VACV D8L | -0.041 | -0.115 | -0.110 | 0.086 | 0.018 | 0.103 | 0.94<br>($p=7.37E-43$) | 1.000 | 0.481<br>($p=1.79E-06$) | 0.278<br>($p=0.0080$) |
| MPXV M1R | 0.145 | -0.038 | -0.130 | 0.033 | 0.195<br>($p=0.0457$) | 0.387<br>($p=5.48E-05$) | 0.531<br>($p=1.22E-07$) | 0.481<br>($p=1.79E-06$) | 1.000 | 0.914<br>($p=6.85E-43$) |
| VACV L1R | 0.103 | -0.046 | -0.094 | 0.232<br>($p=0.0157$) | 0.353<br>($p=0.0002$) | 0.474<br>($p=3.37E-07$) | 0.339<br>($p=0.0012$) | 0.278<br>($p=0.0080$) | 0.914<br>($p=6.85E-43$) | 1.000 |

